# COVID-19 mortality and government response in the Philippines

**DOI:** 10.1101/2025.04.30.25326774

**Authors:** Julius R Migriño, Lorraine Rae S. Martinez

## Abstract

**Objective:** Global response to the COVID-19 pandemic involved the use of public health and social measures (PHSMs) to control and prevent the spread of the virus. The Philippines has adopted these measures during different stages of the pandemic. The objective of this study is to analyze the association of these measures with reported COVID-19 mortality rates.

**Methods:** The study used an analytical cross-sectional document review study design. We used the Oxford Covid-19 Government Response Tracker database for the Philippines from 1 January 2020 to 31 December 2022. The database included 16 policy measures with varying strictness of implementation. We calculated the daily growth of mortality rate based on 6 lag periods representing the lag in effect of the PHSMs at 14, 28, 42, 56, 70 and 84 days. We used multivariate logistic regression models for each lag period to calculate the beta coefficients for each PHSM.

**Results:** Only restrictions on internal movement showed persistent significant negative growth of mortality rates across multiple lag times. Cancelation of public events showed a significant negative growth in mortality rate but only after 84 days. Policies which showed significant positive growth in mortality rate in at least one lag period include protection of elderly people, vaccination policies, restrictions of gathering size, closure of public transport and public information campaign. Policies on school closure, workplace closure, stay-at-home requirements, restriction on international travel, income support, debt/contract relief for households, testing policy, contact tracing and facial coverings showed no statistically significant effect across all lag times.

**Discussion:** Our findings show the implementation of PHSMs in the Philippines is associated with mixed effects on COVID-19 mortality rates, similar to studies from other countries. These suggest that the effectiveness of PHSMs is highly time-and context-specific, and more granular studies on localized policy implementation should be conducted.

## INTRODUCTION

COVID-19 was first detected in Wuhan, China in late 2019, and by March 2020, the World Health Organization declared a COVID-19 pandemic. In response, governments of different countries worldwide have implemented various policies and a wide range of strategies to mitigate the effects of the pandemic.^1^ However, due to a global lack of pandemic response experience, unfamiliarity of a novel disease, and absence of drugs and vaccines against the virus, governments were forced to implement public health and social measures (PHSMs) to control the spread of the virus and the disease.^2^ Categories of PHSMs include travel restrictions, social distancing, and personal protective measures. Furthermore, these also incorporate implementation of lockdowns, creation of quarantine facilities, and effective communication from the ministries of health to the rest of the population.^2^

The Philippines has established the Inter-Agency Task Force for the Management of Emerging Infectious Diseases (IATF-EID) in 2014 as an inter-sectoral collaboration to “establish preparedness and ensure efficient government response… [to] any potential pandemic in the Philippines”. In March 2020, the IATF-EID released the National Action Plan (NAP) Against COVID-19 which outlines the country’s broad strategy to mitigate the effects of the pandemic.^3^ The Philippine government have implemented varying COVID-19 PHSMs at different levels, times and administrative areas throughout the pandemic. The first stricter policy implemented was restriction on international travel, implemented at the end of January 2020 by the Bureau of Quarantine and DOH.^7,8^ By 13 March 2020, through consultation with the IATF-EID, then President Rodrigo Duterte activated Code Red Level 2 and imposed stringent social distancing measures, first in the National Capital Region, but then expanded to the rest of the country at varying levels depending on COVID-19 reports. Such policy variations, or “quarantine protocols”, included the Enhanced Community Quarantine (ECQ), the Modified Enhanced Community Quarantine (MECQ), General Community Quarantine (GCQ), and Modified General Community Quarantine (MCGQ).^4^ Each variation coincided with decreasing levels of a combination of PHSMs such as physical distancing, limitation of gatherings, restrictions in internal and external mobility, workplace restrictions, and closure of public transportation services. In April 2020, the DOH released Administrative Order 2020-0015^9^ which formally provided guidelines for the implementation of specific PHSMs. Due to the heterogeneity of local communities in the country, local government units (LGUs) addressed the health crisis with granular policies specific to their needs. As such, the types and levels of implementation of PHSMs varied across communities.^4^

Assessing and evaluating the effectiveness of public health and social measures is vital for the response plans and prevention of future mortalities. Several studies have determined how PHSMs may have significant effects on the cases and mortality rates of COVID-19.^1,2,5,6^ According to the study done by Haug et al. (2020), the most effective PHSMs are curfews, lockdowns and closures of places like educational institutions, and disallowing small to large gatherings.^6^ Furthermore, no single PHSM can be effective alone yet a combination of PHSMs tailored to the needs of a specific country is maximally effective in the resurgence of COVID-19 cases and mortalities.^6^ However, while some PHSMs may have a mitigating effect on the spread and mortality of COVID-19, some policies may have negative effects on national economies and may exacerbate other socioeconomic problems.^1^ Because of these potential economic and societal tradeoffs, knowing the most effective PHSMs would allow the stakeholders of the healthcare system to implement these elements effectively during a pandemic while minimizing their socioeconomic impacts.

The Oxford Covid-19 Government Response Tracker (OxCGRT) is a real-time, time-series database recording daily government COVID-19 responses across more than 180 countries, including the Philippines, using a standardized set of indicators.^5^ Such indicators are broadly classified as containment and closure policies, economic policies, health system policies, vaccination policies and miscellaneous policies. Each of these classifications is further divided into more specific policies. For instance, containment and closure policies are divided into policies on school closure, workplace closure and restrictions on gathering size, among others. These indicators are coded as ordinal variables, with levels ranging from 0 (no implemented measure) up to 5, with higher values signifying stricter measures for the specific policy.

With the aforementioned approaches and strategies, it is important to gain insight if these policies are effective in limiting COVID-19 mortality in the Philippines. Evaluation of the responses to public health crises such as outbreaks and pandemics is usually underprioritized but important for future preparedness and response measures. The objective of the study was to determine the relationship between the different COVID-19 government policies and COVID-19 mortality using data from OxCGRT.

## METHODS

The study used an analytical cross-sectional document review study design. We used the OxCGRT database (https://github.com/OxCGRT/covid-policy-tracker/blob/master/data/OxCGRT_nat_latest.csv; accessed 17 June 2023), but only limited our data for the Philippines. The OxCGRT record for the Philippines contains 1091 rows representing individual days from 1 January 2020 to 31 December 2022. We used data collected after the country reached a cumulative reported COVID-19 cases of at least 100 based on the COVID-19 Case Tracker from the Department of Health (16 March 2020). We used columns which contained the indices for the 16 policy variables: school closure, workplace closure, cancel public events, restriction on gathering size, close public transport, stay-at-home requirements, restriction on internal movement, restriction on international travel, income support, debt/contact relief for households, public information campaign, testing policy, contact tracing, facial coverings, vaccination policy and protection of elderly people. We also used the column new deaths to represent the daily reported COVID-19 deaths.

Values for the policy variables were re-coded into a binomial scale of 0 = minimal policy (which includes “no policy” as well as less strict policy recommendations) and 1 = stricter policy depending on the policy and their range of indices. The recoding guide is provided in the Supplementary Material. To correct for day-to-day variability in reporting of COVID-19 deaths, we estimated mortality using a 7-day moving average smoothing of new deaths from the OxCGRT database. We then calculated growth of mortality rate *Y*_t+h_ at any time *t* as follows:

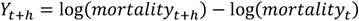

with *h* = {14, 28, 42, 56, 70, 84} representing the lag (in days) of the effects of policies on COVID-19 mortality. Multivariate linear regression models based on the six delay intervals were created using the following specifications:

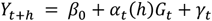

where *α*_*t*_ represents the effect estimate of policy *G*_*t*_, at time *t, β*_*0*_ is the y-intercept and *γ*_*t*_ represents the error term. The augmented Dickey-Fuller test, an assumption test for stationarity for time-series data prior to regression analysis, was done to all growth of mortality rates, and a correlation matrix was used to determine the correlation between different policies. Ordinary least squares regression was used to generate the regression models. To test for autocorrelation, the Breusch–Godfrey test was used on all models, and autocorrelated models were corrected using Cochrane-Orcutt regression. To address heteroskedasticity, the regression models used the Huber/White/sandwich robust estimator.

The results were summarized using unstandardized beta coefficients and semi-robust standard errors from the regression models across all lags. Descriptive statistics and graphs were created using Microsoft Excel while statistical analysis and multivariate linear regression models were generated using JASP (Version 0.17.2.1). An alpha value of 0.05 was used to test for statistical significance. Ethical approval was secured from the San Beda University Research Ethics Board under SBU-REB-2023-004. All data produced in the present study are available upon reasonable request to the authors.

## RESULTS

There have been more than 4 million reported new cases and over 65 000 new deaths related to COVID-19 recorded in the Philippines by OxCGRT during the 1021 days included in the study. The median number of deaths during this time is 30 (IQR 76.25), with most deaths associated with COVID-19 reported between the 2nd quarter of 2021 and the 2nd quarter of 2022. The stricter government policies which were implemented for the longest time include *Public information campaign* (97.45% of recorded days), *Contact tracing* (92.46%) and *Close public transport* (92.07%). Those implemented the shortest include *Stay-at-home requirements* (7.05%), *Cancel public events* (14.79%) and *Debt/contract relief for households* (22.53%). The Philippines implemented a coordinated public information campaign (i.e., information campaigns across different public media) and comprehensive contact tracing (i.e., active contact tracing done for all identified cases) during the majority of the study duration. The timeline of these stricter PHSMs is summarized in Figure 1.

**Fig. 1.**
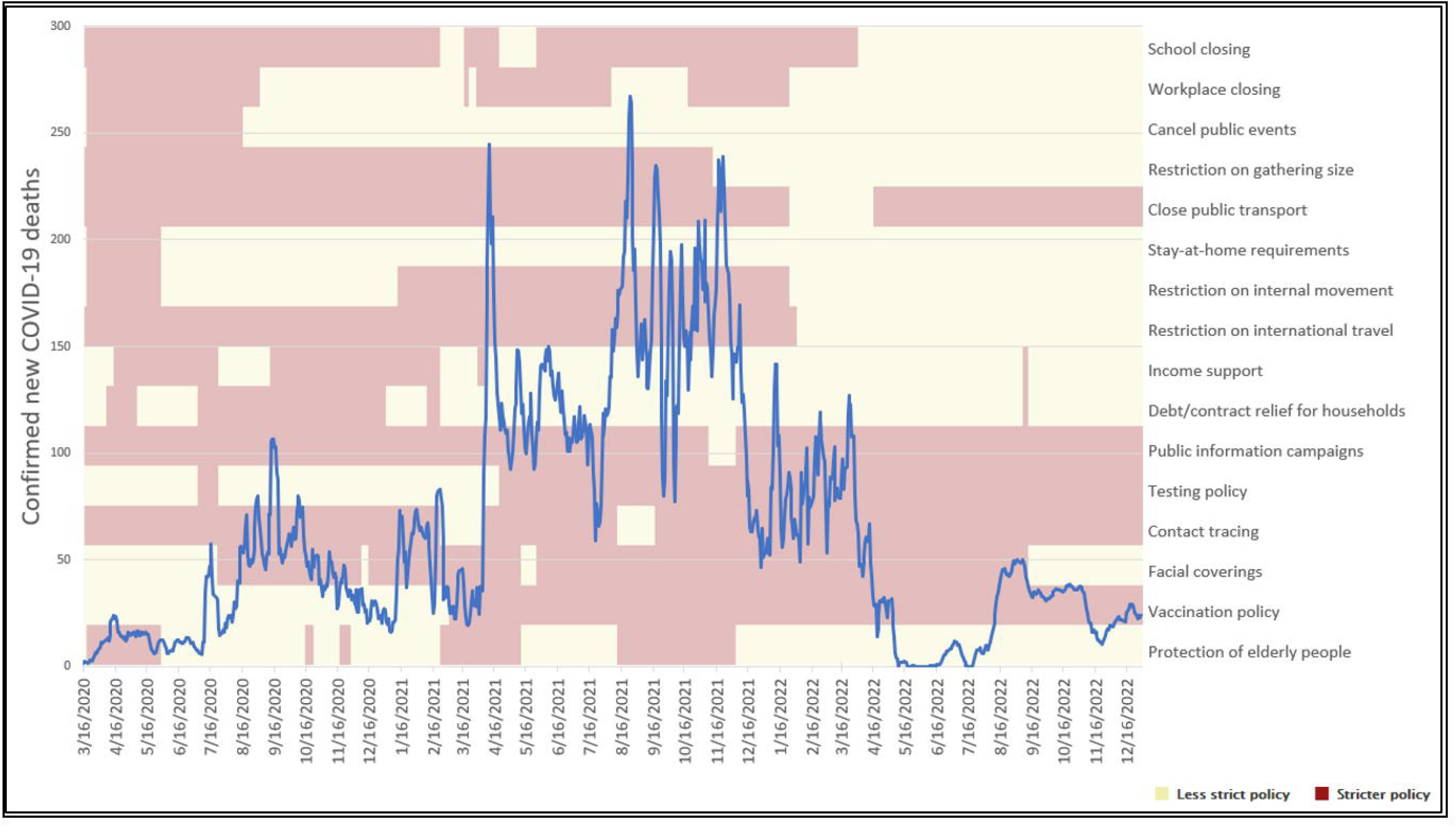
Timeline of stricter COVID-19 government policies and confirmed new COVID-19 deaths since >100 confirmed cases until 31 December 2022

Table 1 shows the effect sizes of the different stricter government policies had on the growth of mortality rate. Among the 16 government policies included in the models, only seven had statistically significant effect sizes in at least one lag time: *Cancel public events, Restrictions of gathering size, Close public transport, Restrictions on internal movement, Public information campaign, Vaccination policy* and *Protection of elderly people* (i.e., recorded policies targeting the protection of elderly people in long term care facilities or in home settings such as restrictions in isolation and limiting visitors). Figure 2 plots the effect sizes of the seven significant policies, along with their 95% confidence intervals.

**Table 1.** Growth of mortality rate based on implementation of stricter government policies by lag time.

| Government policy | Lag |  |  |  |  |  |
| --- | --- | --- | --- | --- | --- | --- |
|  | 14 days | 28 days | 42 days | 56 days | 70 days | 84 days |
| School closure | 0.0212<br>(0.0282) | -0.0097<br>(0.0359) | -0.0316<br>(0.0246) | -0.0097<br>(0.0359) | -0.0928<br>(0.0538) | -0.0315<br>(0.0217) |
| Workplace closure | 0.0341<br>(0.0184) | 0.0063<br>(0.0123) | 0.0016<br>(0.0156) | 0.0063<br>(0.0123) | -0.0019<br>(0.0330) | 0.0290<br>(0.0244) |
| Restrictions on gathering size | 0.0040<br>(0.0461) | -0.0248<br>(0.0501) | 0.0098<br>(0.0601) | -0.0248<br>(0.0501) | 0.4612<br>(0.2206)* | -0.0212<br>(0.0722) |
| Cancel public events | -0.0376<br>(0.0432) | -0.0177<br>(0.0358) | 0.0605<br>(0.0519) | -0.0177<br>(0.0358) | -0.1895<br>(0.1812) | -0.0793<br>(0.0348)* |
| Close public transport | 0.0695<br>(0.0459) | 0.1533<br>(0.0780)* | 0.1525<br>(0.0837) | 0.1533<br>(0.0780) | 0.1596<br>(0.0559)** | -0.0365<br>(0.0566) |
| Stay-at-home requirements | 0.0021<br>(0.0658) | 0.0998<br>(0.0873) | -0.0470<br>(0.0684) | 0.0998<br>(0.0873) | 0.0010<br>(0.1926) | -0.1226<br>(0.0892) |
| Restrictions on internal movement | -0.1095<br>(0.0411)** | -0.1275<br>(0.0493)** | -0.1340<br>(0.0462)** | -0.1275<br>(0.0493)* | -0.1375<br>(0.0367)*** | 0.1226<br>(0.0750) |
| Restriction on international travel | 0.0280<br>(0.0543) | 0.0362<br>(0.0755) | 0.0042<br>(0.0596) | 0.0362<br>(0.0755) | -0.0298<br>(0.0463) | -0.0035<br>(0.0404) |
| Income support | 0.0471<br>(0.0524) | 0.0390<br>(0.0555) | 0.0701<br>(0.0506) | 0.0390<br>(0.0555) | 0.0698<br>(0.0552) | 0.0807<br>(0.0532) |
| Debt/contract relief for households | -0.0346<br>(0.0327) | -0.0117<br>(0.0341) | -0.0170<br>(0.0233) | -0.0117<br>(0.0341) | -0.0425<br>(0.0295) | -0.0335<br>(0.0259) |
| Public information campaign | -0.0064<br>(0.0200) | -0.0439<br>(0.0536) | -0.0652<br>(0.0639) | -0.0439<br>(0.0536) | 0.0625<br>(0.0177)*** | -0.0778<br>(0.0216)*** |
| Testing policy | 0.0033<br>(0.0392) | 0.0174<br>(0.0710) | 0.0023<br>(0.0361) | 0.0174<br>(0.0710) | -0.0202<br>(0.0559) | 0.0214<br>(0.0430) |
| Contact tracing | 0.0091<br>(0.0212) | -0.0245<br>(0.0392) | -0.0483<br>(0.0337) | -0.0245<br>(0.0392) | -0.0132<br>(0.0331) | -0.0429<br>(0.0420) |
| Facial coverings | 0.0131<br>(0.0305) | 0.0486<br>(0.0517) | 0.0513<br>(0.0427) | 0.0486<br>(0.0517) | 0.0709<br>(0.0409) | 0.0107<br>(0.0240) |
| Vaccination policy | 0.0157<br>(0.0440) | 0.0967<br>(0.0459)* | -0.0070<br>(0.0394) | 0.0967<br>(0.0459) | -0.0300<br>(0.0327) | -0.0061<br>(0.0361) |
| Protection of elderly people | 0.0419<br>(0.0177)* | 0.0126<br>(0.0415) | -0.0130<br>(0.0257) | 0.0126<br>(0.0415) | 0.0249<br>(0.0269) | -0.0297<br>(0.0320) |
| Constant | -0.0948<br>(0.0972) | -0.1535<br>(0.1812) | -0.0139<br>(0.2047) | -0.1535<br>(0.1812) | -0.4149<br>(0.2845) | 0.1045<br>(0.2814) |
| R-squared | 0.0044 | 0.0057 | 0.0100 | 0.0057 | 0.0266 | 0.0070 |
| Number of observations | 1006 | 992 | 978 | 992 | 950 | 936 |
| rho | 0.9236 | 0.9554 | 0.967 | 0.9554 | 0.978 | 0.982 |

Note: Semi-robust standard errors are in parentheses: \* $P < 0.05$ , \*\* $P < 0.01$ , \*\*\* $P < 0.001$ .

**Fig. 2.**
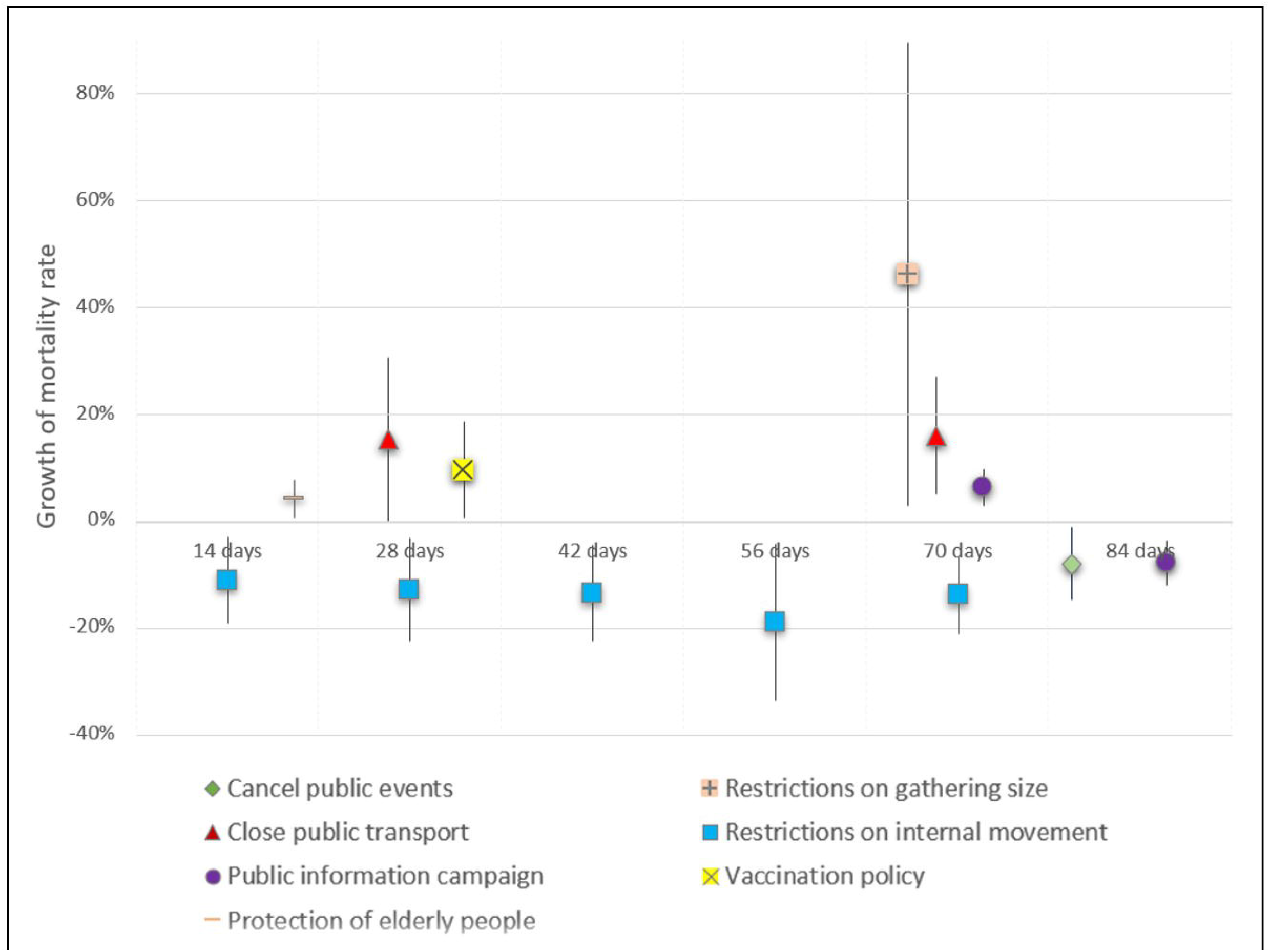
Effects of stricter government policies on the growth of mortality rate by lag time Note: Only values with *P* < 0.05 were included in the graph. Vertical bars represent 95% confidence intervals.

Only *Restrictions on internal movement* showed persistent significant and negative effects on the growth of mortality rates across multiple lag times at 14, 28, 42, 56 and 70 days. The coefficients ranged from - 10.66% at 14 days lag to -18.78% at 56 days lag. This policy had an average effect of a 13.82% decrease in the growth of mortality rate. *Cancel public events* showed a significant negative growth of mortality rate but only at 84 days (-7.93%, 95% CI: -14.76–-1.11%). *Close public transport* showed significant positive growth at 28 (15.51%, 95% CI: 0.07–30.94%) and 70 days (14.71%, 95% CI: 4.36–25.06%). *Protection of elderly people* showed significant positive growth of mortality rate at 14 days (4.19%, 95% CI: 0.68– 7.71%), while *Vaccination policy* showed a significant and positive growth after 28 days (9.67%, 95% CI: 0.47–18.87%). *Restrictions of gathering size* also showed significant positive growth but at 70 days and with a very wide confidence interval (46.12%, 95% CI: 2.83–89.42%). *Public information campaign* showed a significant positive growth of mortality rate at 70 days (6.27%, 95% CI: 3.08–9.46%), but a negative growth at 84 days (-7.78%, 95% CI: -12.03–-3.53%).

The other nine policies (*School closure, Workplace closure, Stay-at-home requirements, Restriction on international travel, Income support, Debt/contract relief for households, Testing policy, Contact tracing, Facial coverings*) showed no statistically significant effect across all lag times.

## DISCUSSION

Our study highlights the persistent and significant effect of restrictions of internal movement on the growth of COVID-19 mortality rates in the Philippines. These policies typically refer to inter-region border checkpoints and closures except for authorized vehicles and personnel. Li et al. (2021) reported that national travel control policies can be effective in decreasing mortality rates as early as 1 week, but these effects were not significant at longer lag times.^10^ Another study did not show substantial reductions in mortality growth from similar policies.^11^ The other policy which showed a significant decrease in mortality growth is cancellation of public events. One study showed a similar result that reported significant negative growth but only at 7 days lag time^2^ compared to our study at 84 days, while another study showed no effect on this policy.^10^ Lastly, one multi-country study found that containment policies may have decreased infection rates but have increased mortality rates.^12^

Several policies in our study showed significant increases in COVID-19 mortality rates: protection of elderly people, vaccination policies, restriction of gathering sizes, and closure of public transportation. One meta-analysis study validates our result, wherein limitations of gatherings increased COVID-19 mortality rates by up to 5.9% based on four studies.^13^ On the other hand, a study by Spiliopoulos (2022) reported that policies on vaccinations are able to decrease both growth of case and mortality rates,^14^ while a study by Clyde et al. (2021) reported that closure of public transportation have a persistently decreasing effect on the growth of mortality rates.^2^ Still, several global studies of OxCGRT data from multiple countries did not find any significant effects of restrictions on gathering size^2^ and elderly protection^11^ on mortality rates. A study in the United States of America reported no significant effect of closure of public transport and public events cancellation policies.^10^ Research on these specific policies is sparse, but some studies agree that vaccine-related policies needed to be coupled with PHSMs for greater efficacy.^15,16^

In our study, one policy showed conflicting effects at different lag times: public information campaigns. One study reported that public information campaigns generally have a significant effect in decreasing case growth rates but not on mortality growth rates across multiple lag times,^10^ which supports our inconsistent findings.

Our study found no significant effects of the other 9 policies on the growth of COVID-19 mortality rates. This is in contrast to previous studies which report positive effects on COVID-19 transmission or mortality by closure of schools^2,13,17^ and businesses,^6,13,17^ stay-at-home orders especially after 1 or 2 months,^2,10,17^ and economic policies such as income or debt support.^12^ Another study however suggests that strict lockdowns and stay-at-home orders may not mitigate COVID-19 transmission and might even be detrimental to the economy and livelihood of low- and middle-income countries.^18^ Seemingly related policies also may have different effects: stricter public testing showed a significant effects on mortality growth rates, but contact tracing did not show these effects.^14^ Using Google and Apple mobility data, Delen et al. (2020) reported that while lockdowns decrease mobility (and thus, transmission) outside, these may increase residential mobility.^19^ Policies on proper and persistent mask wearing as well as on social benefit transfers, on the other hand, has proven to be difficult to enforce due to political, logistical and infrastructure reasons, especially in lower income countries.^20^ and tracking policy effects can be difficult due to inconsistent definitions of “mask mandates”.^13^ Lastly, Mader and Rüttenauer (2022) found that most of these policies did not have any significant COVID-19 mortality effects.^11^

There are several factors that may explain the inconsistent results from our studies. Caldwell et al. (2021) proposes that relatively younger age groups, fewer aged-care facilities and even changes in personal behaviors may have contributed to generally lower COVID-19 health effects in the Philippines.^15^ A study on mobility during the pandemic also noted the importance of business and industry composition in assessing the effects of lockdown measures, with these policies affecting cities with more employment in utilities and hospitality more.^21^ This is invariably important due to the wide differences in industry composition across the multiple regions of the country, and these differences are not tracked by the OxCGRT database. Regardless, multiple studies suggest that combination of PHSMs may be more effective in reducing COVID-19 transmission and mortality.^17,18^ and that earlier interventions generally have a greater impact on growth of death rates.^22^

Our study is limited in that we were not able to analyze PHSM combinations particular to specific localities. This may be a good area for future research and policy considerations since economic analysis simulations in the Philippines predict different economic implications of lockdown policies depending on strictness as well as the implementing region.^23^ Other limitations of our study also include inherent database limitations of OxCGRT such as possible inaccuracies or delays in the coding of policy levels, as well as the use of country-level data despite the heterogeneity in implementation of different policies across the archipelagic country. For instance, most COVID-19-related deaths were recorded in highly dense regions such as the National Capital Region and its neighboring regions.^24^ These limitations may be mitigated by more granular analysis of COVID-19 policies by smaller administrative units, data which is not openly available for analysis. External limitations include the varying transmission and mortality rates of different COVID-19 variants which affected the Philippines^25^ as well as the changing immunologic profile of susceptible populations through time due to vaccines and natural transmission; these factors may affect the growth of mortality rates depending on which timeline is analyzed, and we suggest COVID-19 wave analysis be done in future studies.

Our study finds that the implemented COVID-19 non-pharmacologic intervention policies in the Philippines had varying impacts on the growth of mortality rate. Only *Restrictions on internal movement* was consistently associated with decreased mortality growth rates across multiple lag periods, while cancelation of public events had a small association with decreased mortality growth rates only at a lag period of 84 days. We recommend further studies on the effect of COVID-19 PHSMs within specific administrative units. We also recommend more evidence-based and dynamic implementation of government policies, especially during global or national health events through timely and granular monitoring of such policies.

## Data Availability

All data produced in the present study are available upon reasonable request to the authors.

https://github.com/OxCGRT/covid-policy-tracker/blob/master/data/OxCGRT_nat_latest.csv

## Acknowledgements

The authors would like to thank their families, friends and colleagues for their invaluable support.

## Conflicts of interest

The authors have no conflicts of interest to declare.

## Ethics statement

Ethical approval for the study was given by the San Beda University Research Ethics Board under the study protocol code SBU-REB-2023-004.

## Funding

The study was funded in part by an operational grant from the San Beda University Research and Development Center. The authors had full autonomy in the entire research process, from conceptualization of the proposal to the approval of the final manuscript.

## Notes

### Competing Interest Statement

The authors have declared no competing interest.

