## Supplementary Material for "COVID-19 mortality and government response in the Philippines"

### Appendix A. Recoding manual

1. School closing (0 – no measures; OR recommend closing or open with alterations | 1 – require closing for some levels or categories; OR – require closing all levels)
2. Workplace closing (0 – no measures; OR recommend closing or open with alterations | 1 – require closing for some sectors or categories; OR require closing for all but essential workplaces)
3. Cancel public events (0 – no measures; OR recommend cancelling | 1 – require cancelling)
4. Restrictions on gathering size (0 – no restrictions; OR restrictions on very large gatherings (up to 1000 people) | 1 – restrictions on gatherings below 1000 people)
5. Close public transport (0 – no measures | 1 – recommend (or significantly reduce volume/route/means of transport available); OR require closing or reduce operations)
6. Stay-at-home requirements (0 – no measures; OR recommend not leaving house | 1 – require not leaving house with general or minimal exceptions)
7. Restrictions on internal movement (0 – no measures; OR recommend not to travel between cities/regions | 1 – internal movement restrictions)
8. Restriction on international travel (0 – no restrictions; OR screening arrivals | 1 – quarantine arrivals from some or all regions; OR ban arrivals)
9. Income support (0 – no income support | 1 – government replacing some lost salary)
10. Debt/contract relief for households (0 – no debt/contract relief | 1 – narrow, specific relief; OR broad debt/contract relief)
11. Public information campaign (0 – no COVID-19 public information campaign; OR public officials urging caution | 1 – coordinated public information campaign)
12. Testing policy (0 – no testing policy; OR only those showing COVID-19 symptoms and key workers/admitted to hospital/close contact/from overseas; OR testing anyone with COVID-19 symptoms | 1 – open public testing)
13. Contact tracing (0 – no contact tracing; OR limited contact tracing | 1 – comprehensive contact tracing)
14. Facial coverings (0 – no policy | 1 – recommended or required in any shared/public spaces)
15. Vaccination policy (0 – no availability | 1 – availability for at least one of the following: key workers, elderly, non-elderly vulnerable groups; OR availability for all three plus other groups)
16. Protection of elderly people (0 – no measure; OR recommend protection policies for elderly | 1 – stricter or extensive restrictions for elderly including visitors)
